# Longitudinal changes in infant attention-related brain networks and fearful temperament

**DOI:** 10.1101/2025.01.24.25321085

**Authors:** Courtney A. Filippi, Alice Massera, Jiayin Xing, Hyung G. Park, Emilio Valadez, Jed Elison, Dana Kanel, Daniel S. Pine, Nathan A. Fox, Anderson Winkler

**Affiliations:** Department of Child and Adolescent Psychiatry, New York University, New York, NY 10016; Department of Population Health, New York University, New York, NY 10016; Department of Psychology, University of Southern California, Los Angeles, 90089; Institute of Child Development, Department of Pediatrics, Masonic Institute for the Developing Brain, University of Minnesota, Minneapolis, MN, 55455; Department of Human Development and Quantitative Methodology, University of Maryland, College Park, Maryland, 20742; Emotion and Development Branch, National Institute of Mental Health, Bethesda, Maryland, 20892; Division of Human Genetics, School of Medicine, University of Texas Rio Grande Valley, Brownsville, Texas, 78520

## Abstract

Anxiety disorders are hypothesized to stem in part from altered development of attention-related brain networks. These networks, including the dorsal attention network (DAN), frontal parietal network (FPN), salience network (SN), and default mode network (DMN), are crucial for regulating attention to salient environmental cues. Altered resting-state functional connectivity (rsFC) among these networks in the first months of life relates to fearful temperament, a risk marker for anxiety. Nevertheless, it remains unclear how these networks develop beyond these initial months, particularly in fearful infants. This study characterizes the development of attention-related brain networks over the first two years of life and identifies rsFC changes associated with fearful temperament. Using data from the Baby Connectome Project (from 180 infants across 396 sessions), we analyzed rsFC among the DMN, SN, DAN, and FPN. We examined age-related changes in rsFC across these networks and their relations with fearful temperament at age 2. Results demonstrated age-related decreases in rsFC in DAN – FPN and DMN – SN. Less decrease in DAN – FPN rsFC over time related to greater fear at age 2. This pattern manifested in analyses relating longitudinal trajectories of rsFC and fearful temperament across all available timepoints. Low initial DAN – SN rsFC related to increasing fearfulness over time. This study provides novel insights into the neurodevelopmental origins of fearful temperament.

---

Anxiety disorders are prevalent beginning in adolescence, have an onset in early childhood, and may involve aberrant development of attention-related brain networks (1–5). Attention to salient environmental cues involves the coordination of several large-scale brain networks (e.g., the dorsal attention network [DAN], frontal parietal network [FPN], salience network [SN], and default mode network [DMN]), which develop in infancy (6). Infants at risk for anxiety later in life exhibit altered resting-state functional connectivity (rsFC) among these networks (4,7–11). Yet, it remains unknown how these brain networks change after the first months of life, particularly among infants at risk for anxiety (12). The current study characterizes the development of attention-related brain networks over the first two years of life and identifies functional rsFC changes associated with the fearful temperament, a potent risk marker of anxiety. Fearful temperament, whether measured through observational assessment of behavioral inhibition or through parent-reported fear of novel situations, robustly predicts risk for anxiety— infants exhibiting this trait are more likely to develop anxiety compared to their non-fearful counterparts (13–18). Such risk is particularly pronounced for infants who exhibit high stable fearfulness (19,20). Fearfulness fluctuates over the first year of life, with normative peaks in stranger anxiety occurring around 9 months. By toddlerhood, individual differences in fearfulness appear relatively stable (21–23). Fearful temperament, much like anxiety, involves heightened attention to salient cues, and limited ability to regulate this pattern (24–27).

Several brain networks (i.e., SN, FPN, DMN, DAN) support attentional processes across childhood, adolescence, and adulthood (2,28–33). While the SN directs attention to salient environmental stimuli (34), the FPN and DAN dynamically control the focusing attention to stimuli of interest (35,36). Together, the DAN, FPN, and SN sustain and reorient attention during external attention-demanding cognitive tasks, as needed (35,37). Unlike the SN, FPN, and DAN, the DMN is most active when not engaged in a particular cognitive task (38). For this reason, the DMN is thought to underlie internally environment-orientated attention (39,40). Research links rsFC among these networks to fearful infant temperament (4,7). Specifically, greater rsFC within the SN (7,9,41), less rsFC within the DMN (4,10), and less rsFC between the DAN -DMN, -SN, and - FPN (8) has been related to fearfulness in infancy.

The infant brain undergoes remarkable change over the first year of life (6) and it remains unknown how such changes relate to the development of attention-related brain networks. Addressing this gap requires longitudinal MRI data, but little such data exists (6,11,42–44). Only one report maps how rsFC changes over the first two years of life relate to emotional outcomes. Salzwedel (11) found that greater amygdala- DMN and amygdala– Visual network rsFC and less amygdala- sensorimotor network rsFC (from age 1 to 2) was associated with greater anxiety at age 4. While promising, less than 30 infants in this study provided complete data across all three longitudinal imaging timepoints (i.e., age 0, 1, and 2 years). There is therefore a need for further analyses incorporating larger datasets.

The current study characterizes the development of attention-related brain networks using the Baby Connectome Project (BCP) data (43). The BCP is a large longitudinal MRI study that obtained high-quality rsFC from 180 infants across 396 sessions with most infants providing between 2 and 3 timepoints of longitudinal MRI data. In this study, rsFC among four attention-related networks (i.e., DMN, SAN, DAN and FPN) was quantified, and age-related changes in rsFC were characterized across all network pairs. Next, we evaluated relations between rsFC change and fearful temperament at age 2. We tested whether the rate of change (slope) in rsFC across the first two years of life may be related to parent-report of infant fearful temperament at 2 years. Supporting analyses further probe whether observed patterns persist when considering fearfulness across all available data collection timepoints and when considering initial rsFC. Together, this work aims to provide novel insight into the neurodevelopmental origins of fearful temperament.

## Methods

### Participants

Two hundred and nine full-term infants were scanned between birth and 60 months (UMN site of the BCP; 43). Infants were excluded if they were preterm (gestational age less than 37 weeks), had low birth weight (<2,000 g), contraindication to MRI, if the mother experienced severe pregnancy and/or delivery complications or reported illicit drug use during pregnancy, or if the infant had a first degree relative with a known neurodevelopmental disorder, or had a medical condition affecting development. Infants were also excluded if their parents could not provide informed consent in English or if the child was adopted. All parents/guardians provided informed consent prior to data collection. This study was approved by the Institutional Review Board.

Several infants were excluded for insufficient high-quality MRI data (See Quality Control and Data loss section for details). In total, 180 infants had sufficient high-quality MRI data for inclusion in group-level analyses (see Table 1 for demographics). Of these, 114 infants (63%) contributed high-quality longitudinal data: 59 infants with 2 time-points, 19 infants with 3 time- points, 26 infants with 4 time-points, 9 infants with 5 time-points, and 1 infant with 6 time-points. In total, we utilized MRI data from 396 MRI sessions (*M*MRI sessions per infant= 2.2; SD=1.25, range=1- 6). Infants included in the final analysis did not differ from those excluded in terms of race/ethnicity (*p*>0.51), maternal education (*p*=1) or income (*p*>0.066; See Supplement for more details), and sex (*p*>0.34).

**Table 1.** Demographics for sample of infants with good imaging data.

|  | **BCP** |
| --- | --- |
| **N** | 180 |
| **Sex** |  |
| Female | 91 |
| Male | 89 |
| **Ethnicity/Race** |  |
| Latino/Hispanic and Black | 1 (0.55%) |
| Latino/Hispanic and Asian | 0 (0%) |
| Latino/Hispanic and White | 8 (4.4%) |
| Latino/Hispanic and More than one race | 3 (1.7%) |
| Non-Hispanic and Black | 0 (0%) |
| Non-Hispanic and Asian | 2 (1.1%) |
| Non-Hispanic and White | 137 (76.1%) |
| Non-Hispanic and More than one race | 28 (15.6%) |
| Not_answered/white | 1 (0.55%) |
| **Maternal Education** |  |
| Some High School | 1 (0.55%) |
| High School | 1 (0.55%) |
| Some College | 13 (7.2%) |
| College | 69 (38.3%) |
| Some Graduate School | 11 (6.1%) |
| Graduate School | 80 (44.4%) |
| Not reported | 5 (2.8%) |
| **Income** |  |
| <\$25,000 | 2 (1.1%) |
| \$25,000-\$35,000 | 3 (1.7%) |
| \$35,000-\$50,000 | 6 (3.3%) |
| \$50,000-\$75,000 | 34(18.9%) |
| \$75,000-\$100,000 | 34(18.9%) |
| \$100,000-\$150,000 | 54(30%) |
| \$150,000-\$200,000 | 28(15.6%) |
| >\$200,000 | 18(10%) |
| Not reported | 1 (0.5%) |

### MRI data

#### Acquisition

MRI data were acquired on a 3T Siemens Prisma. At each MRI session, T1-weighted (TR=2,400ms, TE=2.24ms, flip angle=8°, .8 mm isotropic), T2-weighted (TR=3,200ms, TE=564ms, flip angle=variable°, .8 mm isotropic), and resting state (TR=800ms, TE=37ms, flip angle=52°, 2 mm isotropic) scans were obtained. At processing onset, we identified 518 complete MRI sessions. An additional 101 MRI sessions were attempted but resulted in insufficient data for processing (7 infants had insufficient T1-weighted scans, 3 infants were missing T2-weighted scans, 91 were missing resting state data).

#### Preprocessing fMRI

Data from the 518 MRI sessions were submitted to Nibabies (version 21.0.2) for preprocessing. Nibabies, an infant equivalent of fMRIprep, is a standardized workflow that performed B0 inhomogeneity mapping, anatomical data processing, and functional data preprocessing using publicly available methods. These methods leverage field-standard tools including ANTs 2.3.3 (45,46), AFNI (47), FSL 5.0.11 (48), Nilearn 0.8.1 (49), and infant FreeSurfer (50). Anatomical preprocessing used intensity normalization, skull-stripping, tissue segmentation (i.e., gray matter [GM], cerebrospinal fluid [CSF], and white matter [WM]), and spatial normalization to UNC age-specific template and MNI space (51,52). Functional data preprocessing included slice timing correction, estimation of head-motion parameters, spatiotemporal filtering, field map rigid registration to the target echo-planar imaging (EPI) reference run, correction for susceptibility distortions and head-motion, and co-registration to a T1- and T2-weighted reference using boundary-based registration.

Confounds in the preprocessed BOLD time-series were identified and utilized for denoising. These confounds included head-motion estimates and framewise displacement (FD; computed using absolute sum of relative motions; 52), and whole-brain global signal. Frames that exceeded a threshold of 0.25 mm FD or 1.5 standardized root mean square of temporal fMRI signal change at each timepoint (i.e., DVARS) were annotated as motion outliers. The BOLD time-series was then resampled into standard space which produced a preprocessed BOLD time series in MNI Infant space. Non-gridded (surface) re-samplings were performed using mri_vol2surf (infant FreeSurfer).

#### Denoising

Following visual inspection, custom python scripts denoised the data by scrubbing censored frames and regressing out the following nuisance variables: 24 head motion parameters, motion outliers, CSF, WM, global signal, cosines, and pre-steady state outliers. This set of nuisance regressors has been shown to substantially attenuate artifacts in resting state data (54). Low frequency drifts were removed by the inclusion of cosines as nuisance regressors; this method is akin to band-pass filtering (55). Global signal regression was utilized in denoising because it has been shown to effectively remove respiration and motion artifacts (53,56).

#### Quality Control and Data loss

To ensure all processed data was high-quality, quality assessment reports were visually inspected for all 518 MRI sessions. Following visual inspection, 77 MRI sessions (15%) were subsequently deemed poor quality. 27 sessions were outside of the age range of 0-28 months for focal analyses and thus removed. Of the remaining 414 sessions, 18 sessions had fewer than 250 frames of low-motion data. Thus, 396 MRI sessions were deemed high-quality. These 396 MRI sessions came from 180 infants. Averaging across all participants and sessions, our high-quality MRI sample had approximately 11.57 minutes of data (M=867.67 retained frames; SD=370.10 frames; See supplement for distribution). There were no associations between the number of retained frames and age (*p*>.565).

To estimate the relative motion remaining in our high-quality sample, we computed the average FD in retained frames across all infants and across all ages (MFD=.14; SDFD=.017; See Supplemental Table for average by age). While mean FD was low, there was a positive association between age and mean FD (*F*(1) =15.731, *p*<.001; See Supplement Figure 1). Thus, we controlled for mean FD in our models.

### Group Independent Component Analysis (ICA)

To identify the brain networks of interest, we utilized FSL’s Multivariate Exploratory Linear Optimization Decomposition into Independent Components (MELODIC). MELODIC decomposed resting state data into spatial and temporal components. Since our dataset included different numbers of runs per participant and multiple imaging session, we needed a method for ensuring our group map was not biased towards those individuals or timepoints with more available data.

Thus, we constrained the dataset fed to generate the group maps such that every subject that contributed to the group map contributed 2 runs of resting state data at one longitudinal timepoint. This approach ensures that all infants/timepoints contribute to the group network identification process equally. We also ensured that each imaging time point was represented in the group map sample (See Supplement for details). To confirm we tested whether correspondence with the within-network mask did not vary as a function of age. There were no significant age differences for any group map (*p*s>.05). Figure 1 illustrates spatial maps for the attention-related brain networks of interest. Notably, we found two networks that were largely anchored in the PFC and included some limited parietal regions both have been labeled FPN (I & II, respectively). FPN II includes more lateral PFC than FPN I. FPN I encompasses more anterior and medial portions of the PFC. The supplement provides additional details on the anatomical regions highlighted in the group maps.

**Figure 1.**
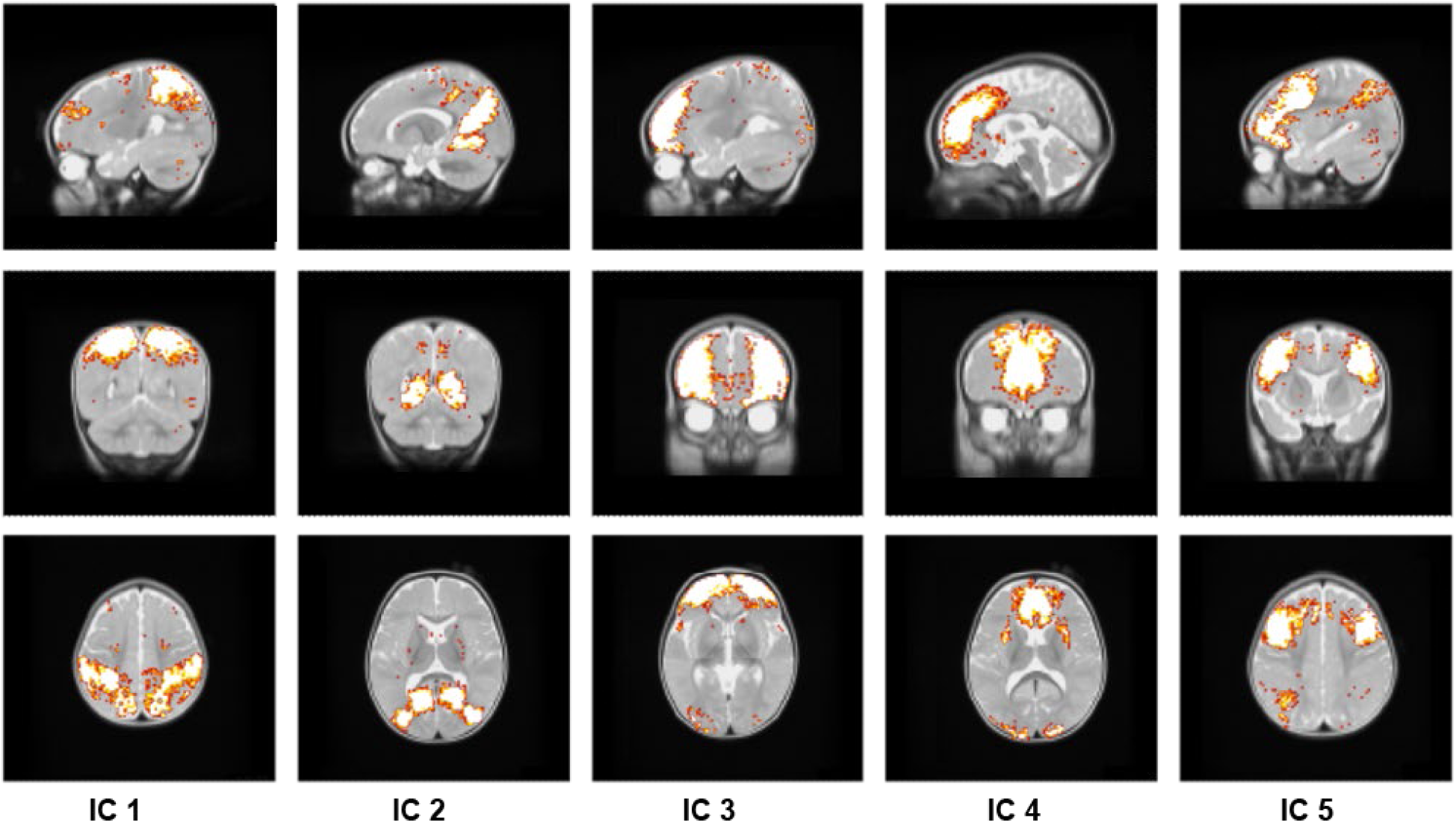
Spatial maps for the 5 independent components selected for this analysis to represent Dorsal attention (IC 1 - DAN); Default Mode (IC 2 - DMN); Salience (IC 4- medial PFC); and Fronto-parietal (IC 3- FPN I; IC 5 – FPN II).

Following MELODIC, we conducted dual regression to extract subject specific time series from the group map. Using the subject-specific time courses as input, we then conducted network modeling using FSLNets functions to compute Pearson correlations between all networks. Fisher’s r-to-z transformation was applied. Figure 2 presents a group-level rsFC matrix with hierarchical clustering. FSLNet’s clustering algorithm detected four clusters. Cluster 1 is comprised of the two FPN components (IC 3 and IC 5). Cluster 2 is comprised of the DAN (IC 1). Cluster 3 is comprised of the SN (IC 4). Cluster 4 is comprised of the DMN (IC 2). In line with other work, we found that: *i*. the FPN is negatively associated with DMN; *ii*. FPN II was negatively associated with SN/VAN; *iii*. DAN/SN/DMN were all negatively correlated with one another (57).

**Figure 2.**
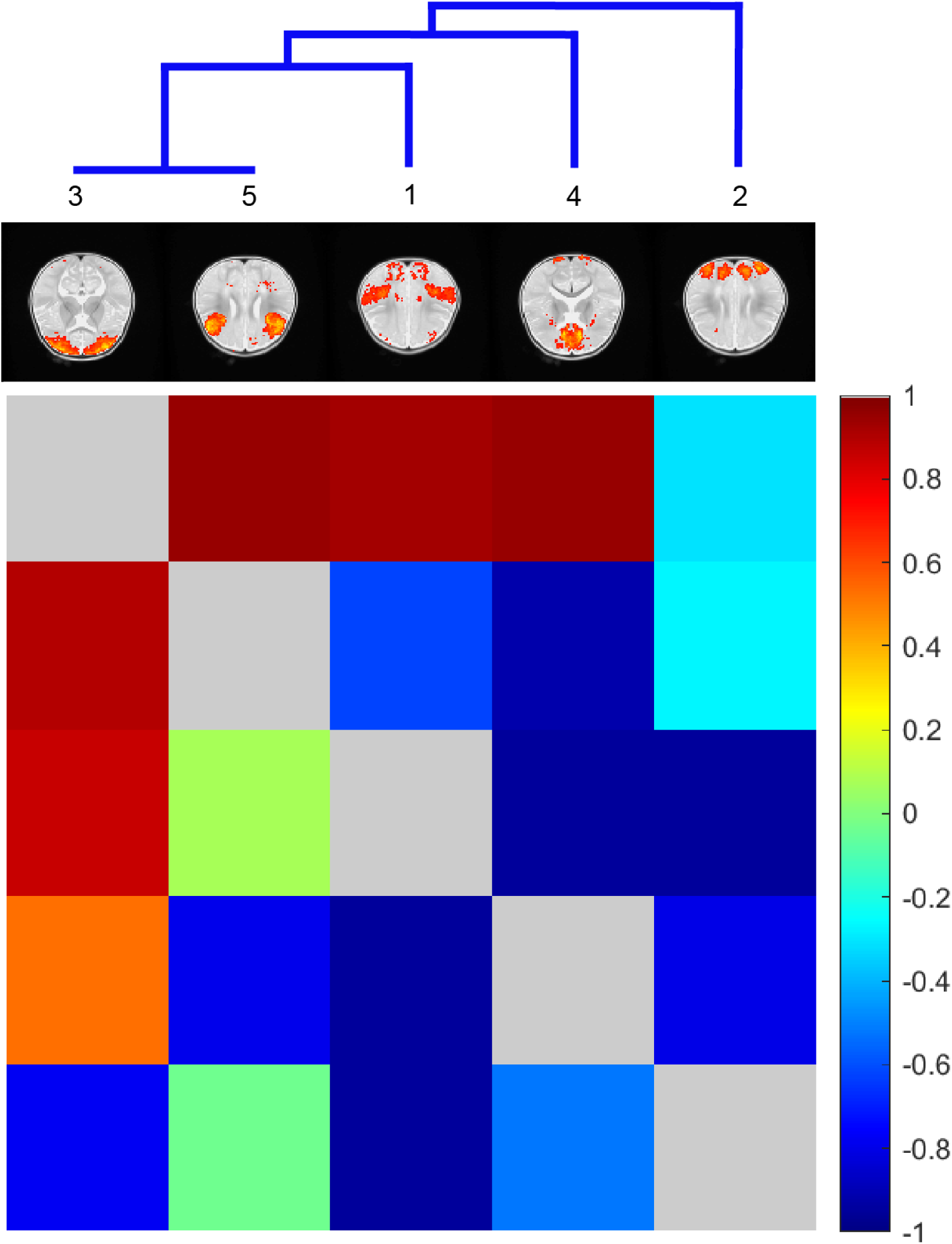
Group-level functional rsFC matrix of resting state data. Full correlations are presented below the diagonal; partial correlations are presented above the diagonal. Four clusters emerged. Cluster components with more similar time courses are closer together.

**Figure 3.**
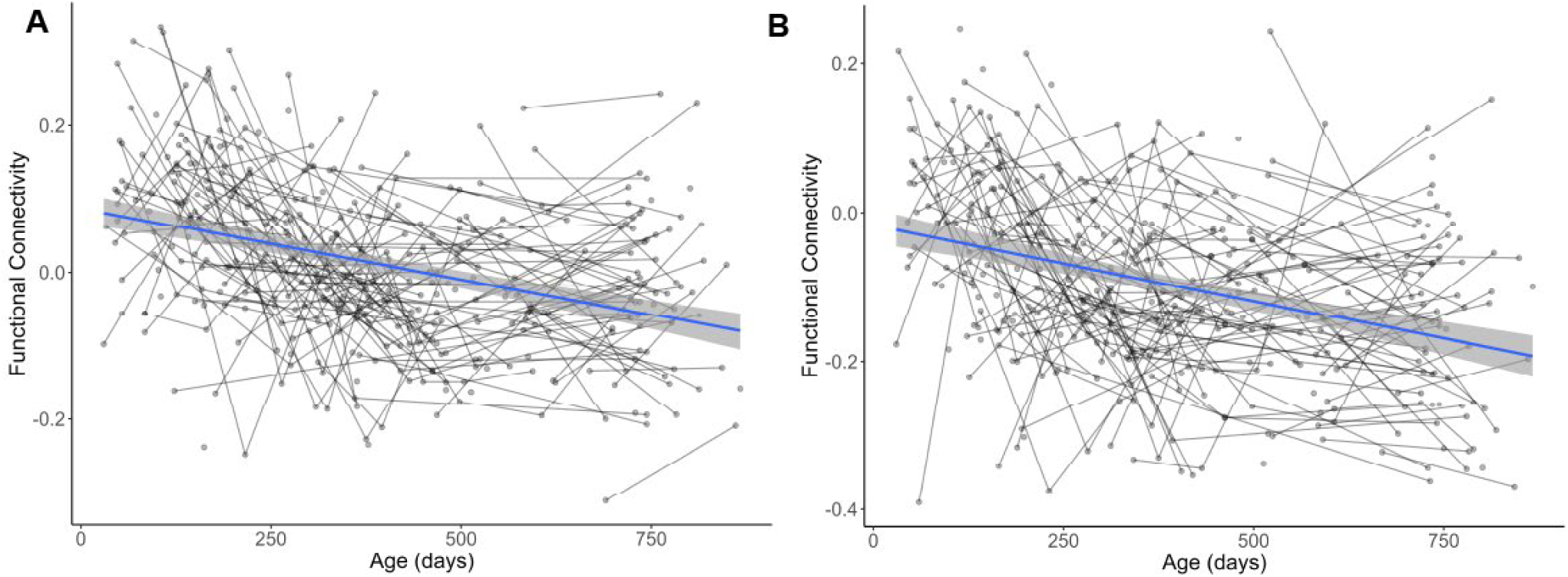
Age-related decreases were observed in DAN – FPN II (A) and DMN – SN (B) rsFC over the first two years of life.

### Parent-Report of Infant Fearful Temperament

Parents reported on infant temperament using the Infant Behavior Questionnaire-Revised (IBQ-R) and the Early Childhood Behavior Questionnaire (ECBQ) (43). All analyses were conducted on the IBQ-R and ECBQ’s Fear subscale which indicates the extent to which the infants exhibit fear in the presence of novelty. The fear sub-scale of the ECBQ is considered equivalent to that of the IBQ, given the identical format and similar question structure, facilitating a seamless age-extension from the IBQ to the ECBQ (58,59). IBQ-R Fear data were collected at 3–5 months (n=68), 6–8 months (n=32), 9–11 months (n=20), and 12–15 months (n=85). ECBQ Fear data were collected at 20–27 months (n=101). A total of 93 subjects provided fearful temperament data at two or more time points, with 57 of them contributing data from at least two time points, including at least one after 20 months of age. For distribution of all available fear data, see the Supplement. ECBQ Fear at 20-27 months (referred to throughout as age 2) was selected as for focal analyses because this was the time point with the largest amount of parent-reported fear data. Exploratory analyses utilized all IBQ and ECBQ data.

### Analytic Approach

The primary study goal was to identify age-related changes in rsFC between the salience (SN), fronto-parietal (FPN I & FPN II), dorsal attention (DAN), and default mode networks (DMN). Thus, we used the average spatial map from the second stage of dual regression to test for age-effects on rsFC. All statistical analyses adjusted for mean FD (framewise displacement, i.e., motion) and sex. Statistical significance was determined using Permutation Analysis of Linear Models (PALM; 58). PALM performs permutation-based inference, providing a robust method for statistical inference without relying on traditional parametric assumptions. All models were run with 500 permutations. The *p*-values were computed after fitting a generalized Pareto distribution to the tail of the permutation distribution (61) to dispense with the need of performing a computationally prohibitive large number of permutations.

The second study goal evaluated whether network changes related to age-2 fearfulness. Thus, for each attention network pair that showed a significant group-level age-related change (DAN-FPN II; DMN-SN), we first extracted subject-level rsFC intercept and slopes using linear mixed effects models, implemented in R v4.4.0 with the lme4 package (62). Specifically, for each network pair, we estimated a linear mixed effects model for the rsFC value, including subject-level random effects for both the intercept and the age term, and extracted the subject-level random intercepts and slopes for subsequent analysis. To model the age effect, we considered both age and log(age), selecting the better representation based on the Akaike Information Criterion (AIC). The model with the lower AIC was reported below (see Supplement for model fit comparison). The intercept and slope were then related to fear through linear regressions, with the subject-level intercept and slope included as a predictor in separate models for fear at age 2. P-values (threshold set at 0.05) for fixed effects were obtained using permutation tests with 10,000 permutations.

Lastly, in an exploratory analysis we examined longitudinal co-trajectories of rsFC and fear across all network pairs and data collection timepoints, using longitudinal structural equation models (SEM). Specifically, for each network pair, we tested the association between network rsFC and fearful temperament in terms of both their initial (intercept) and age-related changes (slope). Given each network pair, this approach jointly estimates the linear trajectory of the corresponding rsFC measures and that of fear, allowing for their longitudinal correlations through shared random effects (63,64). This longitudinal SEM with subject-level random intercepts and slopes is well-suited for accommodating irregular time intervals, mismatch between FC and fear measurements and handling missing data and allowed us to assess the association between changes in each rsFC measure and fear. Given the relatively small sample size with longitudinal assessments of both rsFC and fear, we estimated the models using a Bayesian paradigm for model stability (See Supplement for additional details). This approach allowed us to include 167 subjects with at least one non-missing value for both FC and fear measures. To ensure model estimation stability, we utilized weakly informative prior, which shrink estimates towards the null (i.e., conservative), thereby avoiding overfitting of the longitudinal bivariate associations over time. The significance of FC-fear associations was assessed through the model parameters that capture the bivariate association (represented by the ’Gamma’ parameters, see SI for information) and their credible intervals.

### Results Characterizing attention-related network change over time

On average, significant age-related rsFC decreases manifested in DAN – FPN II (Eta Squared=.13, pFWER<.045) and DMN – SN (Eta Squared=.10, pFWER<.021) (Eta Squared represents the proportion of variance explained by age factor). No other significant changes were observed.

### rsFC slopes associated with 2-year fear

Next, we related both DAN – FPN II and DMN – SN rsFC age-related slopes to fear at age 2 years. Slope of DAN – FPN II rsFC significantly related to fear at 2 (*F*[1] = 4.1993; *β*=2.881, *p* = 0.043; Figure 4). No other significant associations emerged.

**Figure 4.**
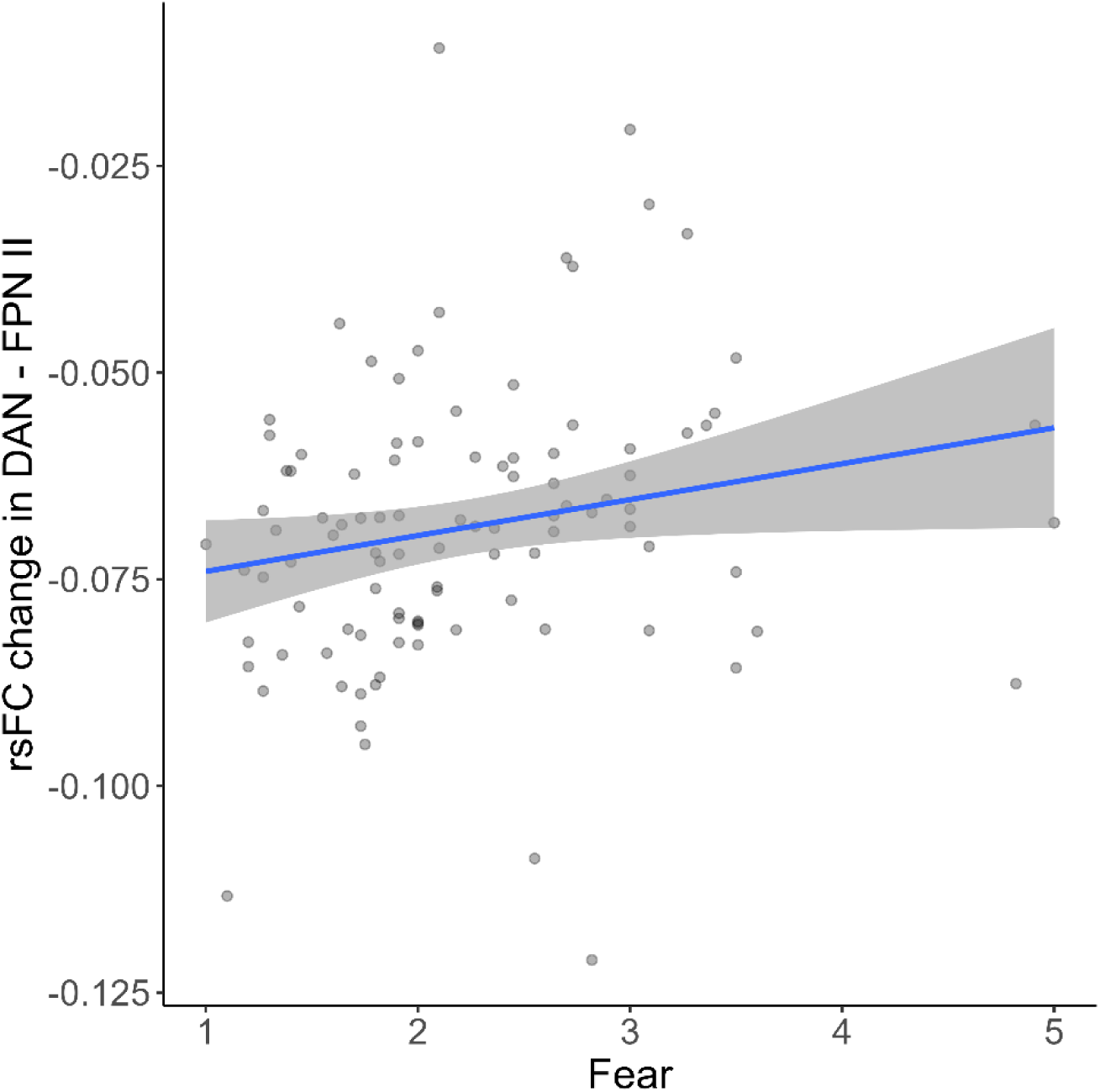
Positive association between rsFC rate of change in DAN – FPN II and fear at 2 years.

### Exploratory analyses: Longitudinal modeling of the association between rsFC and fear over time using all available time points

Lastly, exploratory analyses were conducted to assess whether rsFC trajectories are associated with fear trajectories when considering all available longitudinal MRI and fearful temperament data. The analysis included n=167 subjects with at least one observed value for both rsFC and fear measures (not necessarily observed at the same time points). The trajectory of each of the 10 network pairs was separately related to that of fear measures. Results indicated that the rate of change in fear (slope) had moderate positive associations with: 1) initial DAN-FPN II rsFC value (intercept), with a posterior probability of the corresponding “Gamma” parameter: Pr(Γ_10_>0) = 75%; and 2) the rate of change in DAN-FPN II rsFC value (slope), with a posterior probability of the corresponding parameter: Pr(Γ_11_) = 80% (see Figure 5). Additionally, results demonstrated that the rate of change in fear (slope) had a moderate negative association with initial DAN - SN rsFC value (intercept), with a posterior probability of the corresponding parameter: Pr(Γ_10_<0)=80%. No other notable associations were observed.

**Figure 5.**
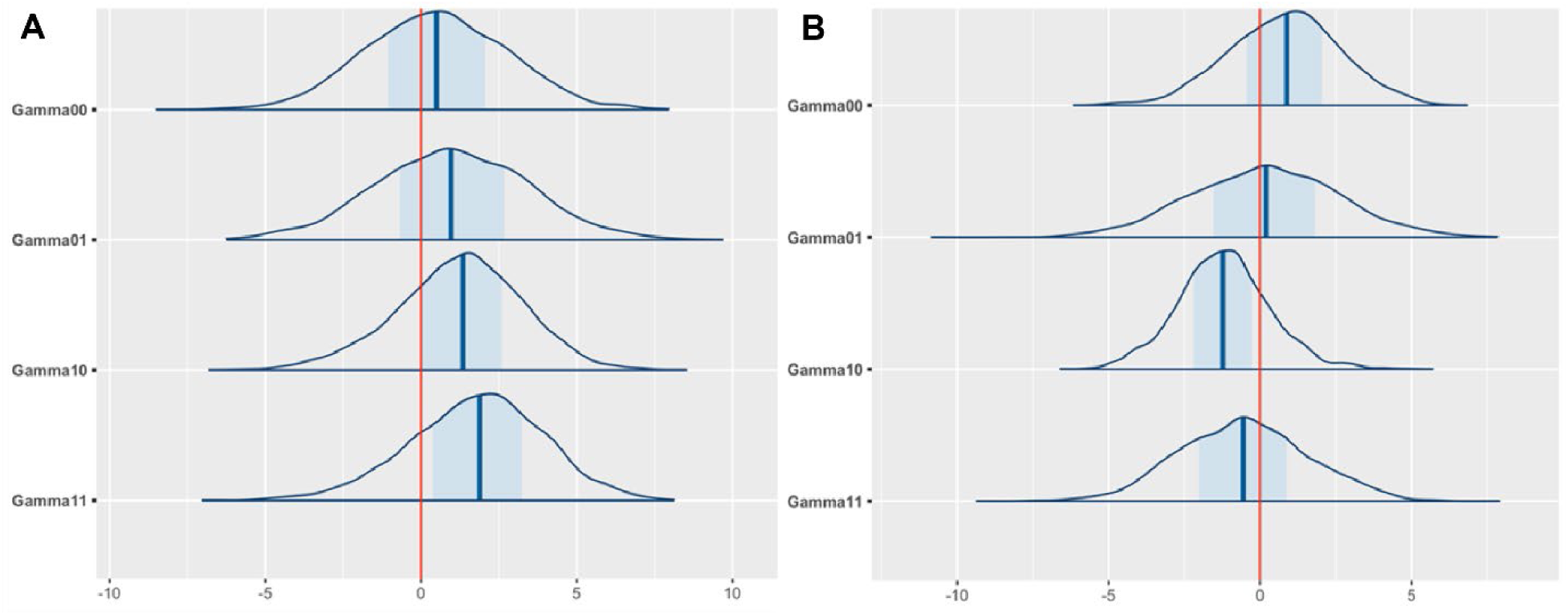
(A) Posterior distributions of ‘Gamma’ parameters for the association between fear and DAN-FPN II rsFC; (B) Posterior distributions of ‘Gamma’ parameters for the association between fear and DAN-SN rsFC. Each panel represents bivariate longitudinal association parameter estimates from the longitudinal SEM for each network pair (DAN-FPN II and DAN-SN): 1) Gamma00 (intercept-intercept) captures the relationship between the fear intercept (initial value) and the FC intercept (initial value); 2) Gamma01 (intercept-slope) captures the relationship between the fear intercept and the FC time slope; 3) Gamma10 (slope-intercept) captures the relationship between the fear time slope and the FC intercept; and 4) Gamma11 (slope-slope) captures the relationship between the fear time slope and the FC time slope. The blue shaded regions represent 50% posterior credible intervals.

## Discussion

This study characterized rsFC change over the first two years of life in attention-related brain networks. Results showed age-related decreases in rsFC in DMN-SN and DAN-FPN. Smaller declines in DAN-FPN rsFC over time were associated with greater fearful temperament at age 2. Supplemental analyses leveraging all the temperament and rsFC data replicated these focal findings within a Bayesian statistical framework. Again, a smaller reduction in DAN-FPN rsFC related to increases in fear over time. These supplemental analyses further demonstrated that greater initial rsFC (i.e., intercept) in DAN-FPN was associated with increasing fearfulness. Additionally, lower initial DAN-SN rsFC was associated with increases in fearfulness. Together, this provides novel evidence that specific changes in attention-related brain networks over the first two years of life (i.e., DAN-FPN rsFC) relate to the development of fearful temperament.

While prior work has found decreased rsFC in DMN-SN and SN-FPN over the first years of life (6), our study is the first to show decreased DAN-FPN rsFC. These findings suggest that attention-related brain networks specialize with development. The higher correlation between activity in these network pairs indicates similar levels of co-activation in early infancy (i.e., less specialization). However, over time this co-activation decreases as each network becomes increasingly well-defined (i.e., more specialized). Neural specialization could support attentional efficiency. This idea is consistent with behavioral findings indicating that over the first years of life, infants develop better attention regulation (using putative top-down processes; 61,62). Our replication of age-related decreases in rsFC between DMN-SN suggests that this finding may be robust to analytic method. In adults, the SN network modulates the balance between DMN and FPN, to achieve both successful detection of external stimuli and its evaluation, based on internal events (32,67). Speculatively, such rsFC changes between the DMN and SN could indicate that, over infancy, increases in regulatory capacity impact the balance between internally and externally oriented attentional control (6,68).

By using a novel Bayesian analytic method which utilizes all available data, we were able to explore how rsFC trajectories relate to the maintenance of fearful temperament over the first two years of life. Our results implicated greater initial DAN-FPN rsFC and lower initial DAN-SN rsFC in increasing fearfulness across the first two years of life. These findings are consistent with previous studies conducted in newborns (4,9), four-month-old infants (8), as well as with work investigating neural correlates of anxiety symptoms in childhood and adulthood (2,68,69). It is well established that the SN and FPN have been implicated in anxiety and early-life risk for developing anxiety (2,70,71), however far less work has outlined the importance of the DAN. The DAN supports selective attention (72,73), and altered DAN rsFC has recently been linked to social anxiety disorders (74). The link between DAN-FPN rsFC in infancy and the maintenance of a fearful phenotype may suggest developmental stability in attention alterations, which could underlie anxiety-related behaviors. This perspective aligns with theoretical frameworks, such as the two-hit model of anxiety, which emphasizes the significance of time-limited experiences in infancy for developmental pathways to psychopathology (75).

In addition to newborn rsFC, we also found that smaller decreases in DAN-FPN rsFC was associated with greater fear. We show this both by relating rsFC slope in the DAN-FPN to fear at age 2 and by using a Bayesian approach. The convergence across these two analytic approaches is promising. By evaluating fearful temperament change in relation to rsFC change over time, these data are the first to demonstrate that changes in attention-related brain networks over infancy (particularly in networks dynamically controlling attention and supporting top-down regulation) may underlie the maintenance of fear over time. This evidence is also in line with several core neurodevelopmental theories of anxiety (24,26). Even so, additional work is needed to understand the extent to which these changes underlie the later development of anxiety symptomatology and to isolate environmental factors that could alter these trajectories.

The current study has several notable strengths. First, we utilized a publicly available dataset from the Baby Connectome Project (BCP). Unlike prior longitudinal studies, this study utilized a cohort-sequential design providing high-quality rsFC data from 180 infants across 396 sessions. In contrast to prior work, which estimates change by leveraging a few developmental anchor points (e.g., 0-3 months, 1 year, 2 years; 6,11), this study’s design facilitated identifying variability across the entire range with nearly all months between 0-2 years. This was possible since most infants contributed between 2 and 3 timepoints of MRI data. Second, our data-driven approach to network identification avoided assuming an adult-like organization for the infant brain. The approach generated an unbiased group map to estimate key infant-brain networks. Third, this paper includes traditional regression analyses using behavioral data at age 2 (i.e., the timepoint when most participants had fearful temperament data). However, we supplemented these analyses using an advanced Bayesian approach, and the consistency of our findings from these supplemental analyses illustrate that our results are robust.

These strengths should be considered in combination with the study’s limitations. First, the correlational nature of the analysis limits interpretations of the causality or directionality of the associations between rsFC changes and the fearful temperament. Future longitudinal studies are needed to further understand how developmental changes in attention networks relate to anxiety outcomes. Second, the study focused exclusively on four key attention-related networks. Thus, it is possible that rsFC among other brain networks (e.g., visual, sensorimotor, auditory; 2,68) may also play a role in the development of fear. Future research is needed to evaluate this possibility. Third, for the measurement of the fearful temperament, we utilized parent-report measures. While these questionnaires provided an efficient assessment of early temperament development in the collection of longitudinal data, we know parent-report and observed behavior are only modestly correlated (76,77). Thus, extensions using observational methods should be conducted to better understand the generalizability of these results. Fourth, while ICA avoids assumptions of adult brain functional organization, it also means that our results are highly dependent on our network identification process. We provide considerable evidence of the regions that make up each network both in the main text and supplement. We caution that our results should be considered in relation to the precise regions captured by our ICA group maps, although the consistency between our findings and other published work using atlas-based approaches may minimize this concern. Additional large-scale longitudinal MRI studies could help determine the reproducibility of these data-driven networks. Finally, the sample reported here is largely non-Hispanic White, highly educated, and greater than 55% of the sample reported an income exceeding $100,000/year. Replicating these findings in more diverse samples is an important direction for future research.

Overall, this study provides novel insights into the development of attention-related brain networks in infancy and how brain change relates to fear in the first years of life. This work is both consistent with neurodevelopmental theories and provides a more nuanced understanding of rsFC changes that may relate to fear. By advancing our understanding of the neural origins of fear, a potent risk-marker for developing anxiety, this work has the potential to inform early identification efforts, and shape future work on the mechanisms that underlie effective early intervention.

## Data Availability

All data produced in the present study are available upon reasonable request to the authors

## Acknowledgements

This research was supported by the National Institute of Mental Health (NIMH) K23MH130751 (PI: Valadez), R00MH125878 (PI: Filippi), R21MH122976 (PI: Fox), R01MH104324 (PI: Elison), U01MH110274 (Co-PI Elison), and intramural NIMH ZIA-MH002782 (PI: Pine). We would also like to acknowledge the families that made this research possible and Isabella Schneider and Hannah Hardiman for their contributions to data cleaning.

