## Supplemental Information for "Longitudinal changes in infant attention-related brain networks and fearful temperament"

### Supplementary Information

#### Participants Methods Extended.

Although the difference was not statistically significant, there was a trend indicating that infants from lower-income families were less likely to have complete data compared to those from higher-income families. Specifically, the mean income for families of excluded subjects was 5.10 (SD = .222), whereas the mean income for families of included subjects was 5.57 (SD = .117). Note that a mean income value of 5 corresponds to an annual income range of $75,000 to $100,000. Future studies should explore this trend further.

#### MRI Methods Extended.

***Evaluating the effect of motion.***

Resting state functional connectivity (rsFC) is highly sensitive to motion. Motion can impact distance-dependent associations in rsFC (1). To ensure that the motion could not explain our effects, we utilized denoising methods that are known to minimize motion-related effects on the BOLD response, such as removing frames with FD > .2 mm, and regressing out global signal (2). Additionally, we evaluated how much retained motion could be impacting our results by evaluating the correlation between retained mean FD and age (See Figure S1). As reported in the main text, there was a small but statistically significant association between mean FD and age. This association appears to be driven by newborns exhibiting lower average mean FD. Newborns small size and common swaddling protocols likely facilitate greater immobilization at data acquisition and thus, greater low-motion data. Although likely related to data collection differences, all subsequent models included mean FD as a covariate to ensure that no within-subject change estimates or group effects could be explained by mean FD.

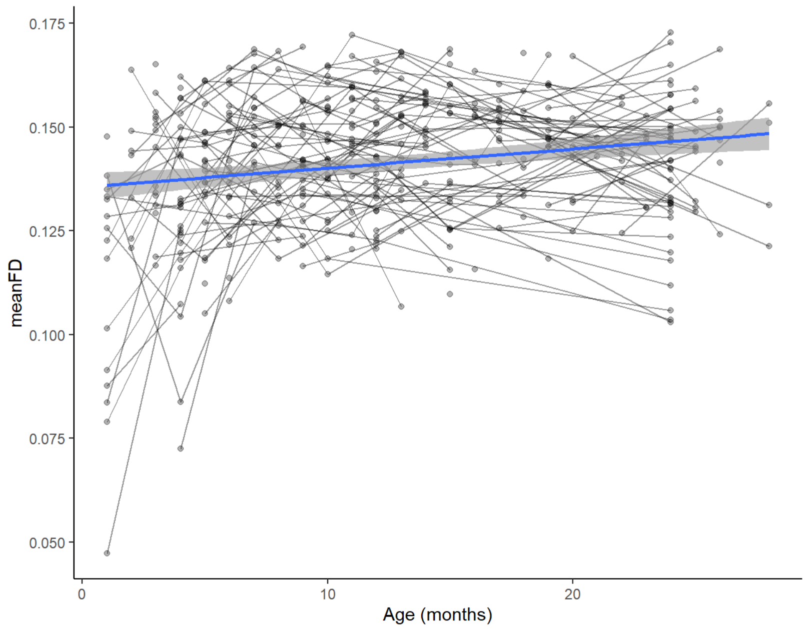


**Figure S1.** Association between mean FD and age.

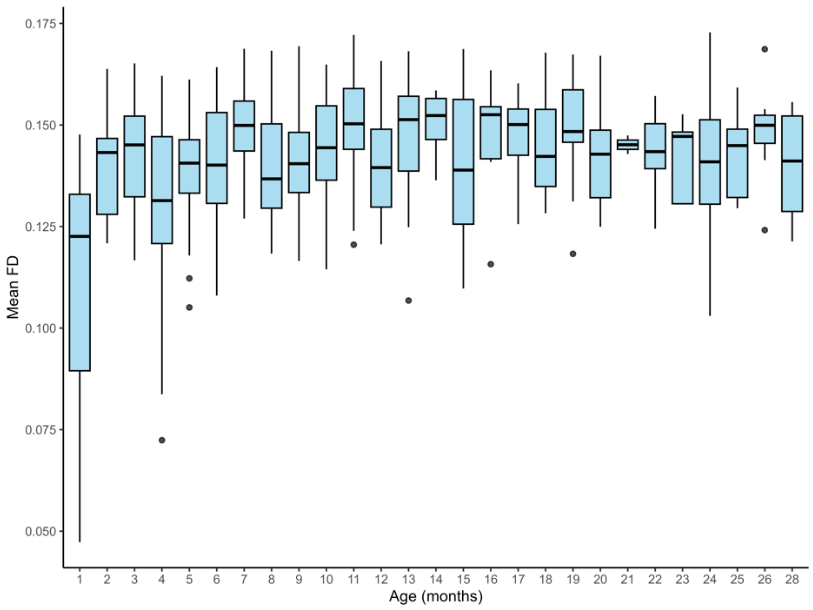


**Figure S2.** Mean (SD) FD by age (months).

***Characterizing high-quality MRI data (after denoising)***

After denoising, most participants contributed 4 runs of resting state data (See Figure S3). As illustrated in Figure S3, very few participants contributed only one run of data and in all cases, these runs had a minimum of 250 frames (see Main Text- Quality control and data loss). All available data on infants between 0-28 months is summarized in Figure S4. While focal analyses examined associations between rsFC and fearfulness at age 2, Bayesian modeling utilized all available data (including earlier parent-reported assessments of fearfulness).

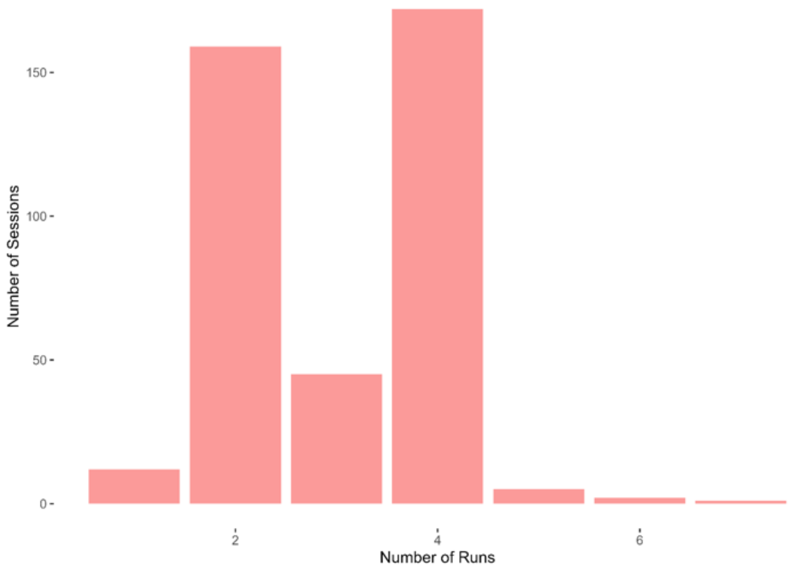


**Figure S3.** Illustrates the total number of runs across all sessions (N=396).

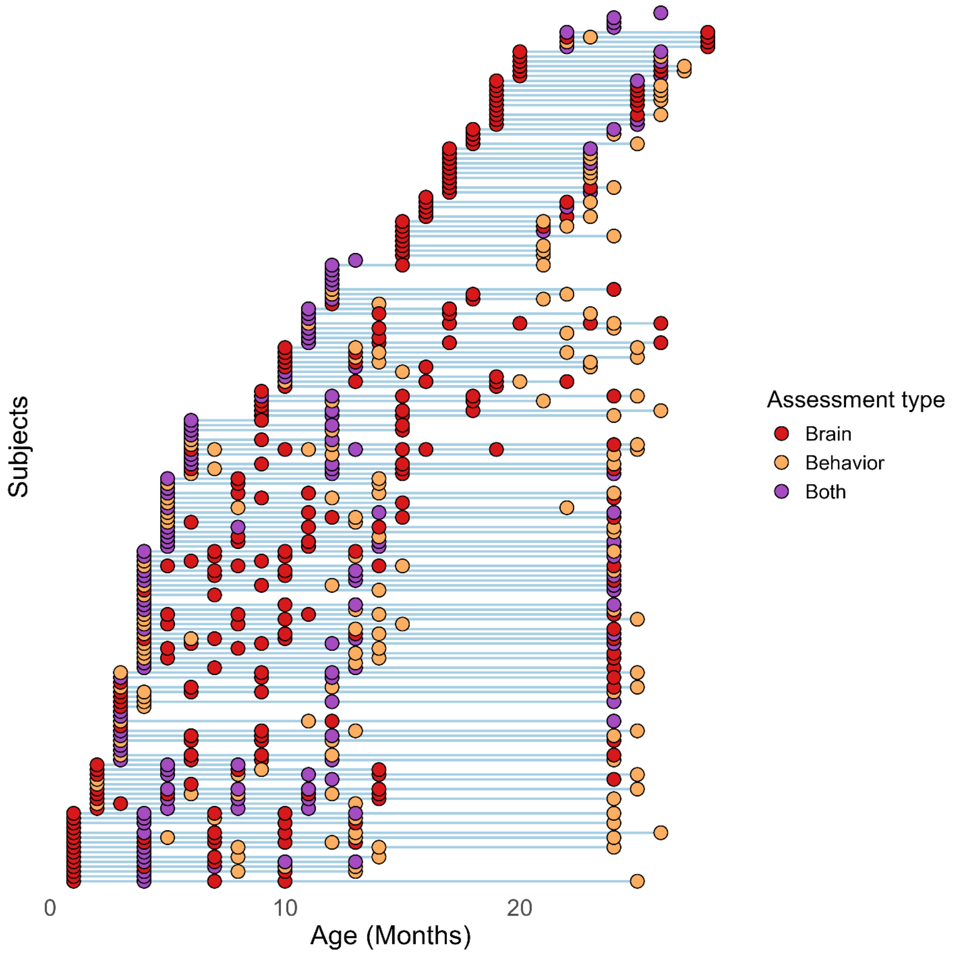


**Figure S4.** All data across all infants, ages and assessment types. Red dots indicate an MRI assessment with high-quality data. Yellow dots indicate behavioral data was available from either the IBQ or ECBQ. Purple dots indicate those instances when at a single timepoint the infant had both MRI and behavioral data available.

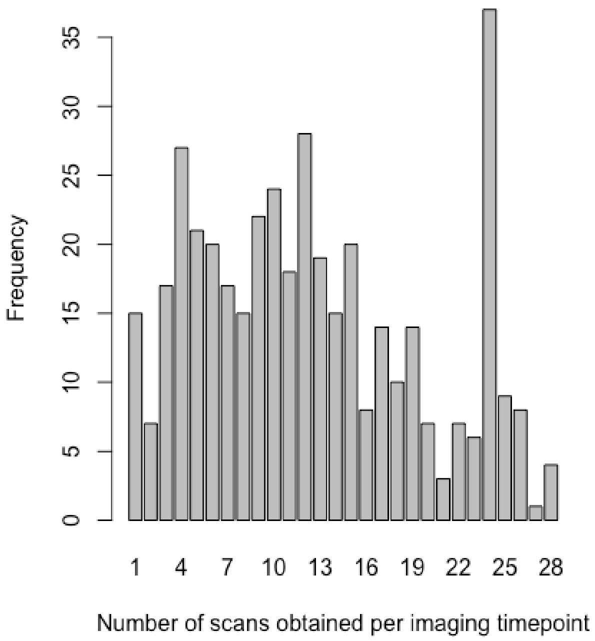


**Figure S5.** Total number of scans obtained at each imaging timepoint.

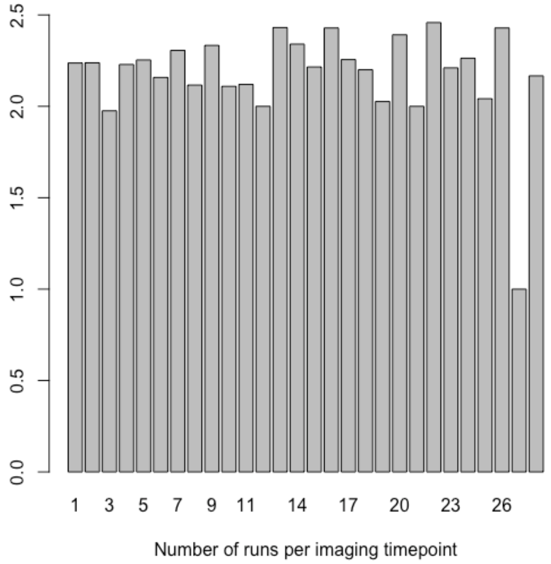


**Figure S6.** Average number of runs obtained for each imaging timepoint.

***Independent Components Analysis (ICA) for network identification.***

To identify networks of interest (as described in the Main Text) we conducted ICA. While Figure S2 illustrates that across all sessions, it was more common for infants to have 4 runs than two runs, these runs were not equally distributed across all imaging timepoints or all participants. Thus, to ensure that across all timepoints most infants were included in the group map, we characterized how many scans were obtained across all imaging timepoints (See Figure S5) and how many runs of data were on average available across each timepoints (See Figure S6). After considering this data and the participant-level contributions, we determined that if the threshold of runs to be contributed was 2, nearly all participants and imaging timepoints would contribute to the group map. Thus, the group map includes a single imaging time point from each participant who had 2 runs of usable data. The timepoint that a particular infant contributed to was determined by assigning infants to the timepoint with the least data first and then balancing the number of infants contributing to each timepoint.

To determine the anatomical regions captured in our group maps, we superimposed the independent components (ICs) over the UNC Infant 1 Atlas (3; See Figure S7 and Table S1).

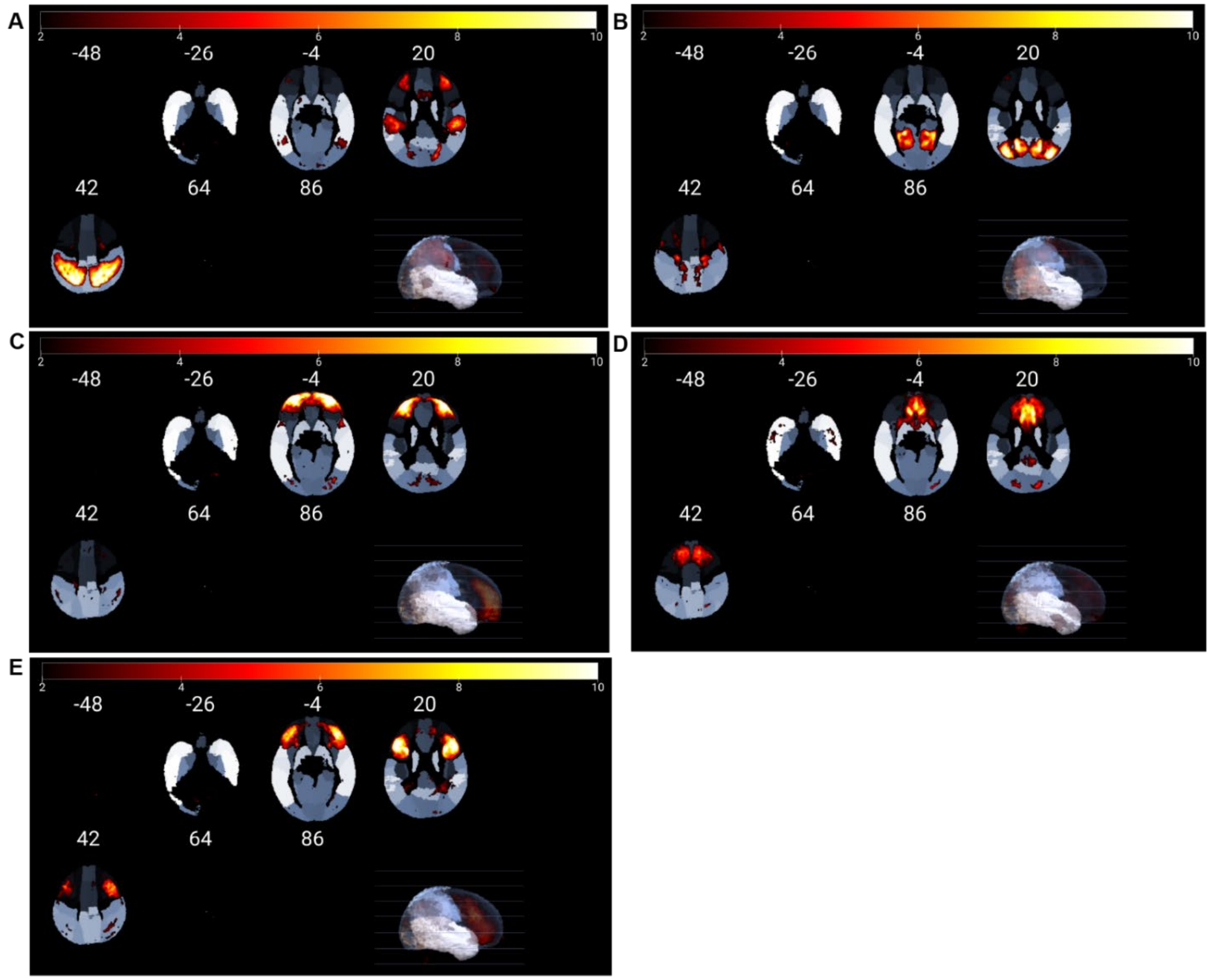


**Figure S7.** Five attention networks ICs superimposed on the UNC Infant 1 Atlas. A) DAN; B) DMN; C) FPN I; D) SN; E) FPN II.

**Table S1. Network labels applied and anatomical regions captured in Figure S7.**

| Network label | Anatomical regions in the group map |
|---|---|
| **IC 1 - Dorsal Attention Network (DAN)** | Precuneus<br>Inferior Parietal Lobule<br>Superior Parietal Gyrus<br>Superior/ Middle Frontal Gyrus<br>Supramarginal Gyrus |
| **IC 2 - Default Mode Network (DMN)** | Middle Occipital Gyrus<br>Lingual Gyrus<br>Precuneus<br>Cuneus<br>Calcarine Cortex<br>Parahippocampal Gyrus<br>Angular Gyrus |
| **IC 3 - Fronto-Parietal Network I (FPN I)** | Middle Frontal Gyrus<br>Temporal Pole Superior<br>Orbitofrontal Cortex<br>Superior Frontal Gryus<br>Cuneus |
| **IC 4 - Salience Network (SN)** | Anterior Cingulate Gyrus<br>Superior Frontal Gyrus<br>Middle Cingulate Gyrus<br>Posterior Cingulate Cortex<br>Superior/Middle Occipital Gyrus<br>Insula |
| **IC 5 - Fronto-Parietal Network II (FPN II)** | Inferior Frontal Gyrus<br>Orbitofrontal<br>Middle Frontal Gyrus<br>Precentral Gyrus<br>Angular Gyrus<br>Inferior Parietal Lobule<br>Heschl Gyrus |

**Behavioral Methods Extended**

The main text's focal analyses use fearful temperament as assessed at age 2 using the ECBQ, a parent-report measure. Additional supplemental analyses utilize a Bayesian approach to make use of all available temperament data. This is possible because the IBQ, a downward extension of the ECBQ, was also obtained. These supplemental analyses examined change in fearfulness over time. Figure S8 depicts average fearfulness over time (with distribution information). Consistent with other work (4–6), we find that fearful temperament increases (normatively) at age 1, then declines at age 2. For this reason, many consider high fearfulness at age 2 to be a robust marker of early-life risk for anxiety.

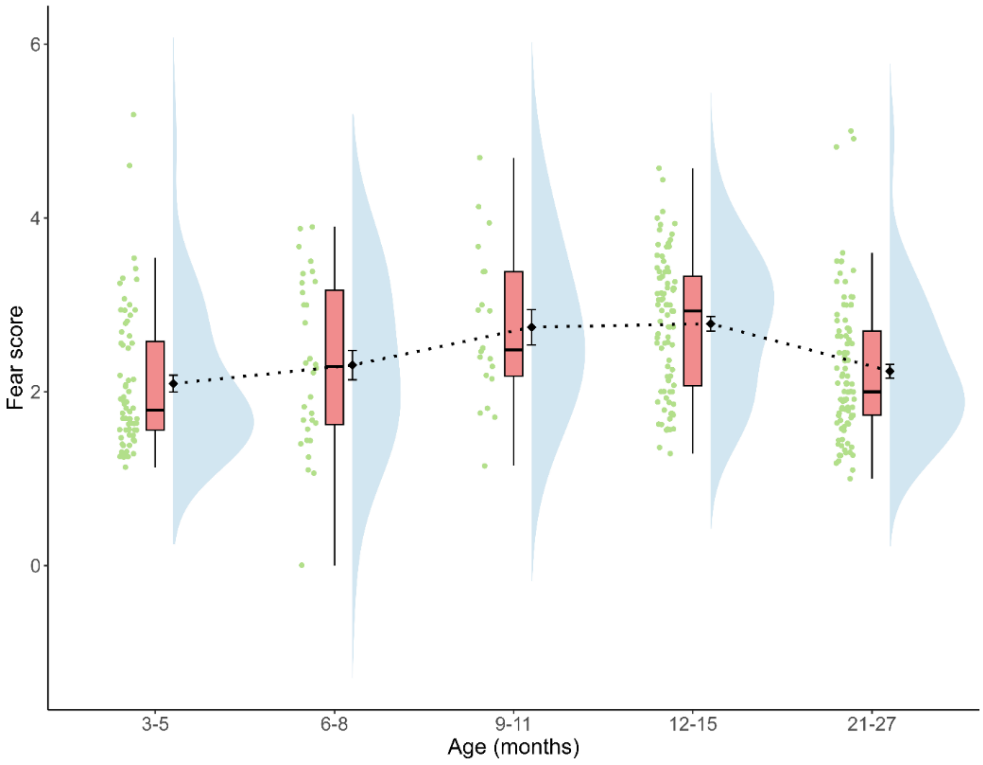


**Figure S8.** Average fearful temperament over the first 2 years of life. Green dots depict subject-level data, Histograms and boxplots provide distributional data. Dotted lines track group-level trends.

### Analytic Approach- Extended

***Focal analyses: Structural equation modeling.***

Longitudinal linear mixed effect modeling was conducted in RStudio version 4.4.0, using the packages: *predictmeans* v1.1.1 (7); *lme4* v1.1-35.3 (8). We modeled network rsFC changes using age and log age. Model fit indices are reported for both models below. Log age was selected based on AIC values (See Table S2).

**Table S2.** Model fit for the linear and the log-linear models.

| Network pair | AIC Age | AIC Log Age | Selected Model |
|---|---|---|---|
| DAN – FPN II | -636.7184643 | -669.324301 | Log-age |
| DMN - SN | -581.9504794 | -603.7966753 | Log-age |

***Exploratory Analyses: Bivariate longitudinal structural equation modeling (SEM).***

The linear growth factor models for connectivity measure for each network pair (denoted as "x") and fearful temperament (denoted as "y") with the corresponding subject-specific intercepts ($\alpha_i^{(x)}$, $\alpha_i^{(y)}$) and subject-specific time slopes (i.e., $\beta_i^{(x)}$, $\beta_i^{(y)}$) are:

$$x_{ij} = \alpha_i^{(x)} + \beta_i^{(x)} \cdot Time_{ij} + \epsilon_{ij}^{(x)}$$

$$y_{ij} = \alpha_i^{(y)} + \beta_i^{(y)} \cdot Time_{ij} + \epsilon_{ij}^{(y)}$$

(we additionally adjusted for mean FD and sex), where the longitudinal growth factors of the connectivity measure (i.e., $\alpha_i^{(x)}$, $\beta_i^{(x)}$) and those of the fear temperament measure (i.e., $\alpha_i^{(y)}$, $\beta_i^{(y)}$), are related through the following structural equation model (SEM):

$$\alpha_i^{(y)} \sim N(\mu_{\alpha_y} + \Gamma_{00}\alpha_i^{(x)} + \Gamma_{01}\beta_i^{(x)},\ \sigma_{\alpha_y}^2)$$
$$\beta_i^{(y)} \sim N(\mu_{\beta_y} + \Gamma_{10}\alpha_i^{(x)} + \Gamma_{11}\beta_i^{(x)},\ \sigma_{\beta_y}^2)$$

and thus, the parameters:

$$\begin{bmatrix} \Gamma_{00} & \Gamma_{01} \\ \Gamma_{10} & \Gamma_{11} \end{bmatrix}$$

of the SEM characterizes the longitudinal association between x and y. The diagram in Figure S9 describes the joint bivariate longitudinal model for "X" and "Y", where the longitudinal evolution ("X1", "X2", …) of the x variable is summarized by "Intercept 1" and "Linear Slope 1", and the longitudinal evolution of the y variable ("Y1", "Y2", …) is summarized by "Intercept 2" and "Linear Slope 2". The association between these growth factors can be summarized by the parameters $\begin{bmatrix} \Gamma_{00} & \Gamma_{01} \\ \Gamma_{10} & \Gamma_{11} \end{bmatrix}$.

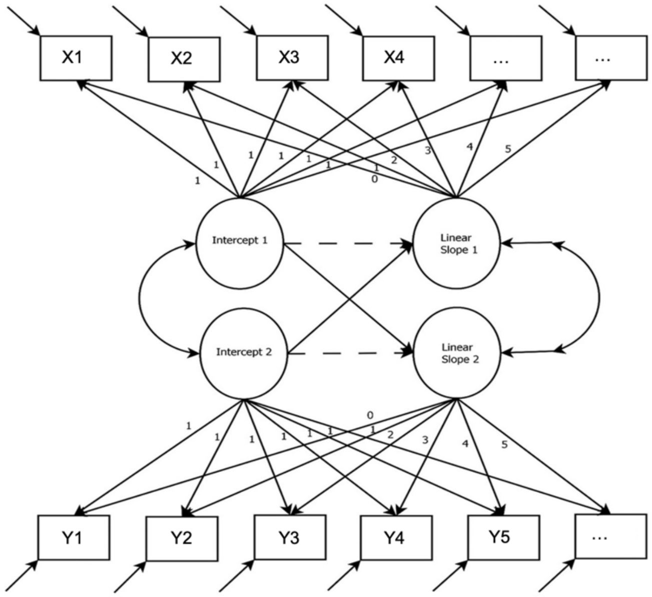


**Figure S9.** Diagram of joint bivariate longitudinal models, where the linear associations between the subject-level intercepts and slopes of the bivariate variables ("X" and "Y") are studied.

Specifically, for the parameters $\begin{bmatrix}\Gamma_{00} & \Gamma_{01}\\ \Gamma_{10} & \Gamma_{11}\end{bmatrix}$, the intercept-intercept relationship $\Gamma_{00}$ captures the relationship between fear (y) intercept and FC (x) intercept; the intercept-slope relationship $\Gamma_{01}$ captures the relationship between fear (y) intercept and FC (x) time slope; the slope-intercept relationship $\Gamma_{10}$ captures the relationship between fear (y) time slope and FC (x) intercept; and the slope-slope relationship $\Gamma_{11}$ captures the relationship between fear (y) time slope and FC (x) time slope.

The model's prior: $\epsilon_{ij}^{(x)} \sim N(0,\ \sigma_x^2)$; $\epsilon_{ij}^{(y)} \sim N(0,\ \sigma_y^2)$;

$\sigma_x \sim Cauchy(0{,}0.1)$; $\sigma_y \sim Cauchy(0{,}1)$; $\alpha_i^{(x)} \sim N(\mu_{\alpha_x},\ \sigma_{\alpha_x}^2)$; $\beta_i^{(x)} \sim N(\mu_{\beta_x},\ \sigma_{\beta_x}^2)$;

$\mu_{\alpha_x} \sim N(0, 1^2)$; $\mu_{\beta_x} \sim N(0, 1^2)$;

$\sigma_{\alpha_x} \sim Cauchy(0{,}0.1)$; $\sigma_{\beta_x} \sim Cauchy(0{,}0.1)$; $\mu_{\alpha_y} \sim N(2.5, 1^2)$; $\mu_{\beta_y} \sim N(0, 1^2)$;

$\sigma_{\alpha_y} \sim Cauchy(0{,}1)$; $\sigma_{\beta_y} \sim Cauchy(0{,}1)$; $\Gamma_{00}, \Gamma_{01}, \Gamma_{10}, \Gamma_{11} \sim N(0, 2.5^2)$.

The priors are specified to scale with the observed data, ensuring they are consistent with the sample's mean and standard deviation. For example, the standard deviation of the "fear" measure was approximately 1, while the standard deviation of the connectivity measure was about 0.1, and to reflect these characteristics, the priors were chosen accordingly. Weakly informative (i.e., conservative) priors were used for the association parameters ($\Gamma_{00}, \Gamma_{01}, \Gamma_{10}, \Gamma_{11}$).
